# Remote self-report and speech-in-noise measures predict clinical audiometric thresholds

**DOI:** 10.1101/2022.05.09.22274843

**Authors:** Lina Motlagh Zadeh, Veronica Brennan, De Wet Swanepoel, Li Lin, David R. Moore

**Affiliations:** Communication Sciences Research Center, Cincinnati Children’s Hospital, Cincinnati, OH 45229, USA; Department of Speech-Language Pathology and Audiology, University of Pretoria, Hatfield 0028, SA; Department of Otolaryngology, College of Medicine, University of Cincinnati, Cincinnati, OH 45267, USA; Manchester Centre for Audiology and Deafness, University of Manchester, Manchester M13 9PL, UK

**Keywords:** tele-audiology, speech perception in noise, hearing loss, digits-in-noise, SSQ

## Abstract

Developments in smartphone technology and the COVID-19 pandemic have highlighted the feasibility and need for remote, but reliable hearing tests. Previous studies used remote testing but did not directly compare results in the same listeners with standard lab or clinic testing. This study investigated reliability of remote, self-administered digits-in-noise (remote-DIN) compared with lab-based, supervised (lab-DIN) testing. Predictive validity was further examined in relation to a commonly used self-report, Speech, Spatial, and Qualities of Hearing (SSQ-12), and lab-based, pure tone audiometry. DIN speech reception thresholds (SRTs) of adults (18-64 y/o) with normal-hearing (NH, N=16) and hearing loss (HL, N=18), were measured using English-language digits (0-9), binaurally presented as triplets in one of four speech-shaped noise maskers (broadband, low-pass filtered at 2, 4, 8 kHz) and two digit phases (diotic, antiphasic). High, significant intraclass correlation coefficients indicated strong internal consistency of remote-DIN SRTs, which also correlated significantly with lab-DIN SRTs. There was no significant mean difference between remote- and lab-DIN on any tests. NH listeners had significantly higher SSQ scores, and remote- and lab-DIN SRTs than listeners with HL. All versions of remote-DIN SRTs correlated significantly with pure-tone-average (PTA), with the 2-kHz filtered test the best predictor, explaining 50% of variance in PTA. SSQ total score also significantly and independently predicted PTA (17% of variance) and all test versions of the remote-DIN, except the antiphasic BB test. This study shows that remote SSQ-12 and remote-DIN are sensitive tools for capturing important aspects of auditory function.

## Introduction

The World Health Organization (WHO) estimates that the number of individuals with disabling hearing loss (i.e., threshold > 35 dB HL for the better ear) has been growing and is expected to increase to 2.5 billion by 2050 (WHO 2021). Unaddressed hearing loss in adults not only affects communication, psychosocial well-being and quality of life, but also has a substantial socio-economic impact (Olusanya et al. 2014). The global annual cost of unaddressed hearing loss is estimated at 980 billion US dollars (WHO, 2021). Late-diagnosed or untreated hearing loss also has detrimental effects on children’s and youth’s communication and cognitive skills, emotional well-being, and academic success (Moeller et al. 2007; Punch et al. 2004; Warner-Czyz et al. 2015). Early detection and treatment of hearing loss is thus both care- and cost-effective in preserving hearing and quality of life for affected people (Davis et al. 2007; Karpa et al. 2010; Maharani et al., 2018).

Existing in-person audiological service delivery models are unable to address the global burden of hearing loss. Low- and middle-income countries (LMICs) experience barriers accessing specialized hearing care services due to factors including shortage of audiological professionals, equipment costs and centralized services (Swanepoel & Clark, 2019; Swanepoel & Hall, 2010). Other factors such as transportation costs and accessibility, limited mobility and poor health also restrict patients’ access to necessary hearing healthcare services, even in high income countries (Powell et al. 2019; Coco et al. 2016; Swanepoel & Hall, 2010). The COVID-19 pandemic has further exposed the need for remote, valid hearing health care options.

Advances in mobile technology and global connectivity through the internet have allowed development of sensitized web- and app-based screening and diagnostic tools that can improve assessment of auditory function in several ways. For example, smartphone deliverable variants of the digits-in-noise test (DIN) can reliably detect and differentiate sensorineural and conductive hearing loss (Smits et al. 2005; Potgieter et al. 2018; Swanepoel et al. 2019; Motlagh Zadeh et al. 2020; De Sousa et al. 2018, 2020, 2021). DIN is a relatively undemanding speech-in-noise test that measures speech recognition abilities objectively, reliably and quickly, in addition to having a strong correlation with audiometric thresholds (Smits et al. 2006; Ozimek et al. 2009; Leensen et al. 2011; Vlaming et al. 2014; Folmer et al. 2017; Motlagh Zadeh et al. 2019, 2020). Moreover, unlike audiometry, DIN testing (speech reception threshold; SRT) is accurate across different devices and headphone types without the requirement of calibration (Vlaming et al., 2014; Potgieter et al. 2016).

DIN testing typically presents digits to listeners against a simultaneous background of broad-bandwidth, speech-shaped noise (Smits et al, 2004). SRT is the usual outcome measure; the speech (digit) signal-to-noise ratio at which three successive digits are all correctly recognized on 50% of presentations (Vlaming et al. 2014). During the past 10 years, the efficacy and sensitivity of the standard DIN test have been substantially increased using various test modifications such as low-pass filtering of the noise (Vlaming et al. 2014; Motlagh Zadeh et al. 2019; Motlagh Zadeh et al. 2020) and antiphasic presentation of the digits (De Sousa et al. 2018; De Sousa et al. 2020). For example, De Sousa et al (2020) showed that presenting digits that are phase inverted (antiphasic) between the ears, while leaving the masking noise interaurally in-phase, significantly improves sensitivity of the DIN test (area under the receiver operating characteristic curve, AUROC = 0.94). If diotic presentation is also used, the DIN differentiates conductive or unilateral from bilateral sensorineural hearing loss (De Sousa et al, 2021). Using low-pass filtering of the noise, Motlagh Zadeh et al (2020) sensitized the DIN test (92% sensitivity and 90% specificity) to high-frequency hearing loss as low as 20 dB HL from 2-14 kHz.

The ability to deliver hearing tests outside the clinic has been long recognized as a potential major benefit of the DIN. In this study we examined the predictive validity for pure-tone audiometry of self-administered DIN versions delivered to the same participants in the lab and at home. Previous studies have also used remote or self-administered DIN versions (Smits et al, 2004 and 2006; Folmer et al, 2017; Swanepoel et al, 2019; De Sousa et al. 2021) but, to our knowledge, no study has directly compared results in the same listeners with standard, rigorous lab or clinic testing.

Self-report is playing an increasing role in hearing assessment. The currently recognized complexity of hearing mechanisms, both within and outside the conventionally defined auditory system (Moore, 2018), reduces the likelihood that one, or even a small number of tests can capture the full experience of hearing. Self-report can provide an overview of the listener experience in different circumstances and can also comment more widely on the usability and effectiveness of test procedures, and how hearing loss impacts on everyday life (‘participation’). However, validation studies of self-report scales have generally been based on other scales and pure-tone audiometric thresholds (Humes et al, 2019). Further examination of the relationship between self-report scales, speech-in-noise and other supra-threshold measures is needed. In this study, we relate and contrast pure-tone audiometry and DIN test with an e-version of the 12 item Speech, Spatial, and Qualities of hearing scale (SSQ) completed at home by participants with either normal or impaired hearing. The original, 49 item SSQ measures self-reported ability for spatial hearing, sound segregation, and hearing speech in a variety of real-life contexts (Gatehouse et al. 2004). Noble and colleagues (2013) showed that SSQ-12 provides similar results to the original SSQ.

According to the Global System for Mobile Communications Association (GSMA; 2020), 3.8 billion people were mobile internet users by the end of 2019. This number increased by 250 million compared to 2018. Importantly, 90% of new users were from LMICs. Validating reliable and sensitive hearing self-assessment tools thus has potential to increase access to hearing healthcare and subsequently improve quality of life for large global populations.

Overall, this study aimed to validate the effectiveness of the remote, self-administered DIN test and SSQ-12. Specific aims of the study were to 1) measure test-retest reliability of remote-DIN SRTs in both normal hearing (NH) participants and those with hearing loss (HL), 2) compare DIN mean SRTs in remote- and lab-based conditions using broadband and filtered noise, and phasic and antiphasic digits, 3) compare SSQ scores with remote-DIN SRTs and, 4) determine the relationship between DIN and SSQ scores with pure tone average (PTA) hearing thresholds and, thus, the relative ability of DIN and SSQ to detect HL.

## Materials and methods

### Participants

Two groups of adult participants, 16 with normal hearing (NH; mean = 34.2 y/o, SD = 13.7) and 18 with hearing loss (HL; mean = 49.1 y/o, SD = 13.8), mostly college graduates (64%; M= 34.2 y/o), were recruited from the Cincinnati Children’s Hospital Medical Center (CCHMC) employee intranet and the local community via information flyers. Flyers provided a brief description of the study and asked for adults 18 to 65 years old to contact our study staff if they have normal hearing, or either a diagnosis of hearing loss or if they suspect having a hearing loss. Interested participants (N= 42) received an initial screening questionnaire that enquired about their demographic information and history of ear and hearing disorders. Participants who met the hearing status criteria (described below) and age range (N= 40) were contacted by the study staff to schedule an assessment date. These participants received routine clinical assessment, including otoscopy, tympanometry, and pure tone audiometry to rule out conductive pathology and determine hearing thresholds across the standard frequency range (0.25 – 8 kHz). Participants who did not meet the inclusion criteria for hearing status (i.e., either normal hearing, or sensorineural hearing loss at standard frequency range AND normal status of outer and middle ears) were excluded from the study (N= 6).

All eligible participants completed the SSQ-12 remotely via the CCHMC Research Electronic Data Capture (REDCap) URL and performed both remote and in-lab versions of DIN tests. Due to scheduling and participants’ availability during the pandemic, 18 of 34 participants (7 NH and 11 HL) completed the lab testing (audiometry and DIN) 3 to 6 months before remote testing (SSQ and DIN). The remaining 16 participants (9 NH and 7 HL) did the remote testing 2 to 4 weeks before lab testing. Consent was given per guidelines set by the CCHMC Institutional Review Board.

### Audiological testing

Pure tone audiometry was performed during lab testing using an Interacoustics Equinox 2.0 audiometer, calibrated to ANSI 3.6 (2010) standards. Thresholds from 0.25 to 8 kHz were obtained from participants in a double-walled sound booth meeting criteria of ANSI S3.1. Sennheiser HDA300 circumaural headphones were used to obtain air conduction thresholds. HL was defined as audiometric thresholds > 20 dB HL in either ear at any frequency from 0.25 to 8 kHz.

### Digits in noise (DIN) testing

#### Stimuli and SRT determination

The digits 0-9 were recorded and homogenized for equal intelligibility based on the method described by Motlagh Zadeh et al. (2019). Each presentation trial consisted of 3.25 s of masking noise in which three different digits were presented with an interval of 175 ms between digits. The noise started and stopped 100 ms before and after each triplet presentation and was fixed at 65 dB SPL. The average length of each digit was 0.66 s. In Diotic mode, triplets were presented binaurally in-phase in one of 4 different noise maskers: broadband (BB) and three low-pass filtered (cutoff at 2, 4, 8 kHz) speech-shaped noises. We also tested binaurally antiphasic triplets in the BB noise version (De Sousa et al., 2020).

BB noise was constructed by summing the long-term average frequency spectrum across all digits. Low-pass noise maskers were constructed using a 10th-order Butterworth low-pass filter with three different cutoff frequencies (2, 4, 8 kHz), summed with a 15 dB attenuated version of the original BB noise. A one-down, one-up adaptive procedure was used to obtain SRTs. A correct response was defined as responding to all three digits correctly. The final SRT was calculated as the average signal-to-noise ratios (SNR) of the final 19 of 23 total trials. For detailed information regarding recordings, homogenization, and noise masker construction refer to the Supplementary Information link provided by Motlagh-Zadeh et al (2019).

#### Remote implementation

The DIN test was provided via the hearDigits® research website (hearX Group). The fixed order of DIN testing was as follows: 1-4) Diotic digits in BB, 2 kHz, 4 kHz, and 8 kHz filtered noise; 5) Antiphasic digits in BB noise; 6) repeat Diotic digits in BB noise to ascertain short-term test-retest reliability. Testing took about 30 minutes to complete.

Participants were instructed to use whatever headphones (or earbuds) they had available with any device (PC/smartphone/tablet) that had a compatible web browser (Chrome, Firefox or Safari). Studies have shown that DIN tests are generally insensitive to headphone quality (Swanepoel et al. 2019; Vlaming et al, 2014). Each remote generated .csv file contained detailed test information including: participant identification number, name, date of birth, test mode (Diotic or Antiphasic), noise version (BB or low-pass filtered at 2, 4, 8 kHz), and trial by trial presented triplets, SNR and responses.

#### Lab implementation

The lab DIN-SRTs were also obtained via the hearDigits® research website and followed the same order of DIN testing as the remote procedure. The lab-DIN test was administered using Sennheiser 215 headphones connected to an iPad in a double-walled sound booth meeting criteria of ANSI S3.1. No test-retest was done for the lab testing since the reliability of lab procedures has already been established (Motlagh-Zadeh et al, 2020).

### SSQ Scale

An e-version (administered via REDCap) of the SSQ-12 (Noble et al, 2013) with 9 subscales (Table 2) was given to participants at the time of remote testing. As specified, participants were instructed to respond to prompts by moving a virtual slider that ran from 0 to 10, where 10 meant they were perfectly able to do or experience what was described in the question, and where 0 meant they were quite unable to do or experience what was described in the question. The average score across all subscales was calculated and used as a measure of self-reported speech perception in noise ability.

### Analysis

Initially, univariate analysis was conducted to explore the impact of individual variables (SSQ score and DIN-SRTs) on the outcome (better ear PTA) via Pearson correlation and t-test. Variables of at least marginal significance (*p* < 0.1) in the univariate analysis were considered as potential predictors for a subsequent multiple regression. To assess the relative importance of individual predictors, multicollinearity was measured by variance inflation factors (VIF). Variables with VIF greater than 5 were removed sequentially. Stepwise regression was performed to select the most parsimonious model. The final parsimonious linear regression model included two significant variables (Hearing Status Group (NH vs HL), *p* = 0.02; and 2 kHz low-pass filtered DIN, *p* = 0.0004) which explained 70% of the variance in BE-PTA. Their VIFs were less than 1.70, indicating no obvious collinearity. All data were analyzed in SAS v9.4 (The SAS Institute, Cary, NC) with two-sided significance levels of 0.05.

## Results

All 34 participants who met the inclusion criteria completed home- and lab-based testing successfully.

### Audiometry

Figure 1a shows the hearing thresholds for both NH and HL groups. Half the participants in the HL group (n = 9) had PTA < 20 dB (average of all frequencies in both ears, Fig. 1b). Two of 18 participants in the HL group (dashed gray lines, Fig. 1a) had asymmetric loss (interaural difference ≥15 dB at three contiguous frequencies; AAO-HNS, 1993 and 1994). The HL group was thus heterogeneous in both degree and symmetry of their hearing loss. However, when the entire analysis was repeated following removal of the two participants having asymmetric loss, no difference in the main results or conclusions was obtained. The presented data thus include those participants.

**Figure 1.**
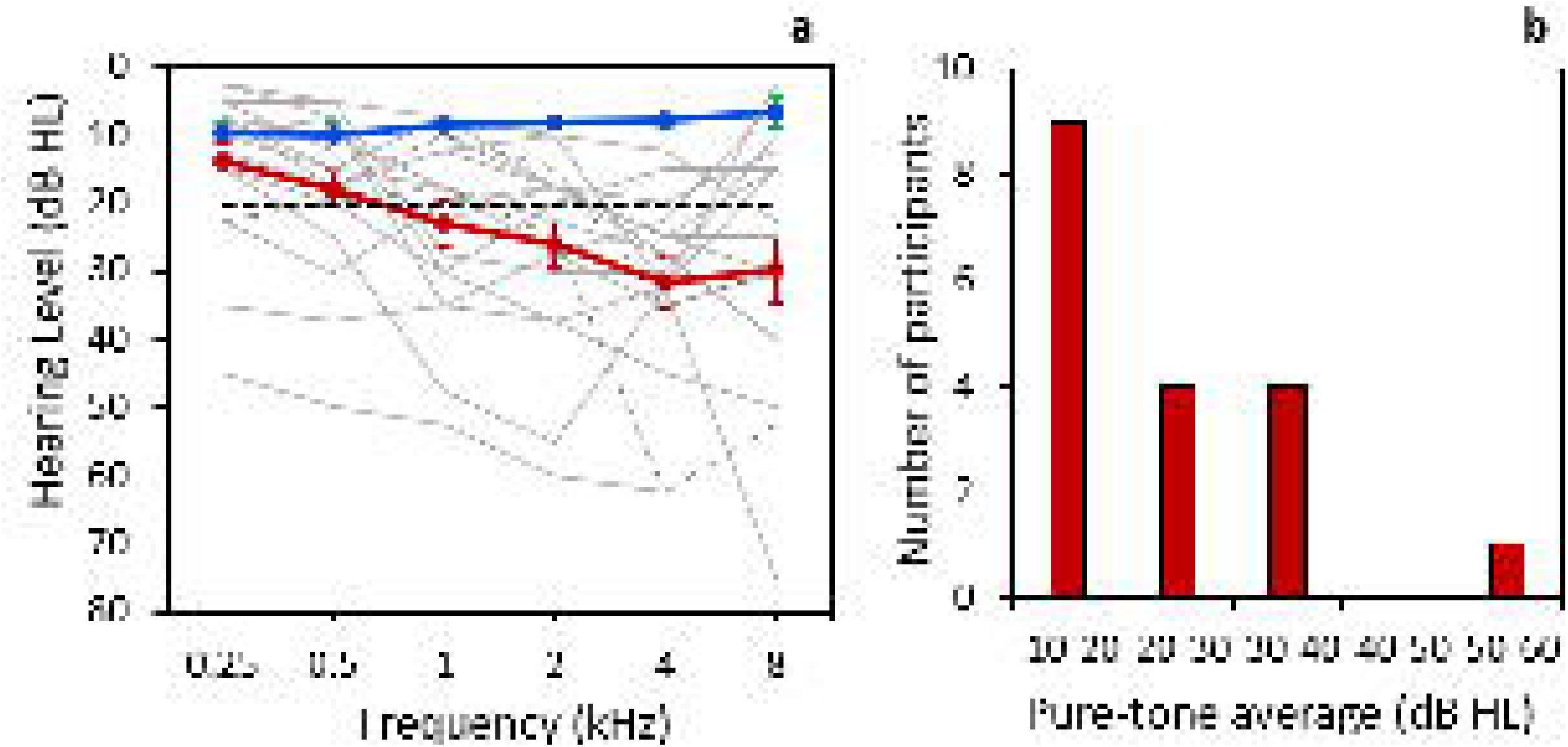
Audiometric thresholds. **a)** Average thresholds and standard errors expressed in hearing level for the normal hearing (NH, blue) and the hearing loss (HL, red) groups. Gray lines show HL individual audiograms and the high variability in their hearing thresholds. Dashed gray lines show individuals with asymmetric hearing loss. Black dashed horizontal line shows the level of clinical normal hearing sensitivity (≤ 20 dB HL). **b)** Distribution of pure tone average across all frequencies in both ears in the HL group (N = 18). For each range of PTA, only the lower measure is inclusive (e.g., 10-20 = 10-19.99).

### Reliability of remote DIN test

Test-retest reliability was measured for Diotic BB Remote-DIN in all participants. A high and significant intraclass correlation coefficient (ICC=0.80, *p* < 0.0001) indicated strong internal consistency and reliability of the remote-DIN test (Fig. 2). Reliable results were found separately for both NH (ICC=0.58, *p* < 0.01) and HL (ICC=0.80, *p* < 0.0001) groups.

**Figure 2.**
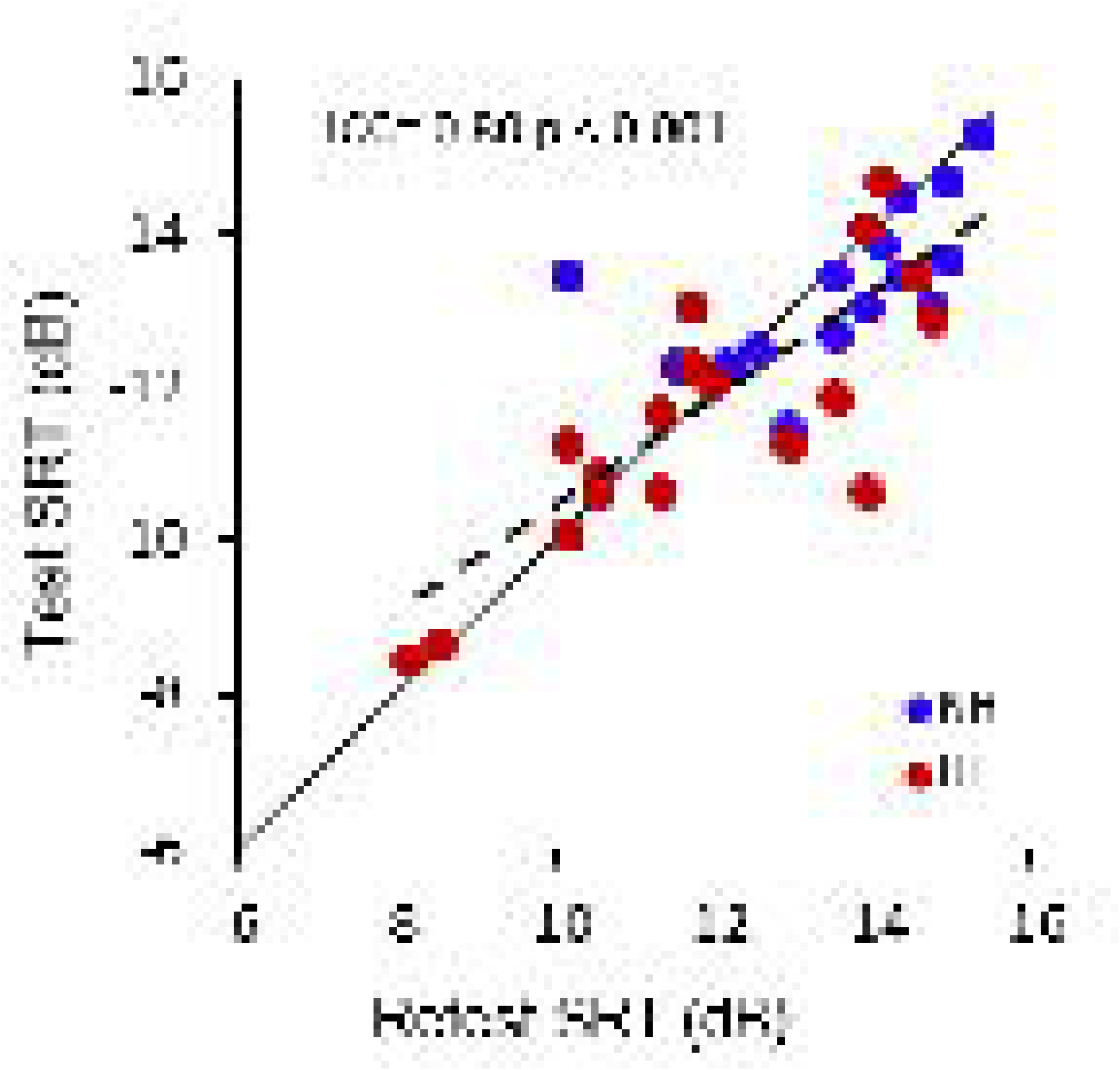
Remote-DIN test-retest reliability. Speech reception threshold (SRTs) for the diotic BB test in normal hearing (NH) and hearing loss (HL) groups. Black solid line shows Test = Retest and the dashed line shows the least-square regression line for NH and HL groups combined.

### Remote- vs Lab-DIN test

Table 1 shows the mean DIN-SRTs for remote and lab test conditions in the NH, HL, and both groups combined. Mean remote SRTs did not differ significantly from mean lab SRTs in any test version. Across all participants, lab- and remote-SRTs correlated significantly in all test versions (Fig. 3, black regression lines). Diotic-BB was the only test version in which the lab- and remote-SRTs were significantly correlated in both groups of participants (NH and HL). Regression lines in Figure 3 suggest a trend toward better SRTs in the lab for listeners with HL. However, there were outliers in both environments in this group and all differences between environments were non-significant.

**Figure 3.**
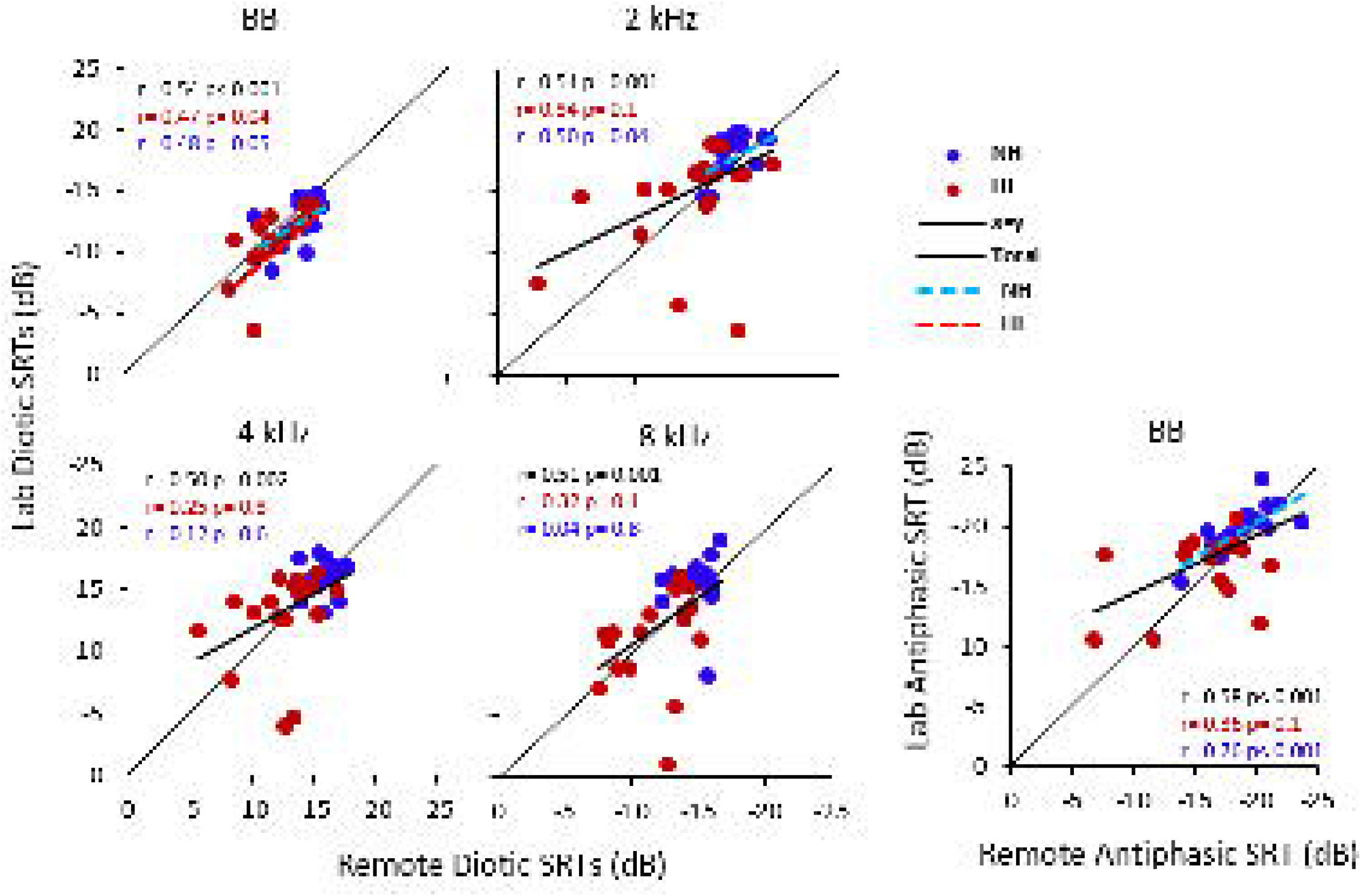
Comparison between lab- and remote-tested SRTs. Lab and remote speech reception thresholds (SRTs) for the diotic broadband (BB) and low-pass filtered (2, 4, 8 kHz cutoffs) DIN tests and for the antiphasic BB DIN test in the normal hearing (NH) and hearing loss (HL) groups. A more negative SRT indicates more sensitive hearing. Least-square regression lines indicate significant correlations among the Total sample (bold) and within each group (dotted).

**Table 1.** Mean (SD) SRT for lab- and remote-DIN tests for each noise version. Only the first.

| Participants | Diotic |  |  |  |  |  |  |  | Antiphasic |  |
| --- | --- | --- | --- | --- | --- | --- | --- | --- | --- | --- |
|  | BB |  | 2 kHz |  | 4 kHz |  | 8 kHz |  | BB |  |
|  | Lab | Remote | Lab | Remote | Lab | Remote | Lab | Remote | Lab | Remote |
| **NH** | -12.4<br>(1.8) | -13.6<br>(1.4) | -18.0<br>(1.7) | -17.6<br>(1.4) | -15.9<br>(1.3) | -16.1<br>(1.1) | -15.4<br>(2.3) | -14.9<br>(1.3) | -19.7<br>(2) | -18.9<br>(2.5) |
| **HL** | -10.2<br>(4) | -11.9<br>(2) | -14.0<br>(4.3) | -14.1<br>(4.2) | -12.6<br>(3.6) | -12.4<br>(2.7) | -11.1<br>(4) | -11.8<br>(2.5) | -16.5<br>(3) | -16.0<br>(4) |
| **Total** | -11.3<br>(3.2) | -12.7<br>(2) | -15.9<br>(3.8) | -15.8<br>(3.6) | -14.1<br>(3.2) | -14.2<br>(2.8) | -13.1<br>(3.8) | -13.3<br>(2.6) | -18.0<br>(3) | -17.3<br>(3.5) |

obtained Remote BB data (Fig. 2) were included.
*Normal Hearing (NH); Hearing Loss (HL); Speech reception threshold (SRT); BB: Broadband noise; 2 kHz: 2 kHz low-pass filter noise; 4 kHz: 4 kHz low-pass filter noise; 8 kHz: 8 kHz low-pass filter noise*

There were two outliers who had substantially poorer lab-DIN SRTs compared to remote-DIN SRTs (Fig. 3). Removing those outliers improved the correlation between lab- and remote-DIN SRTs, but we saw no specific reason for excluding them.

The subgroup (N= 18; 7 NH and 11 HL) who completed the lab testing (audiometry and DIN) 3 to 6 months before remote testing (SSQ and DIN) had significantly (*p*= 0.02) higher mean DIN-SRTs than the subgroup (N=16; 9 NH and 7 HL) who did the remote testing 2 to 4 weeks before lab testing. We did not find a significant mean difference between test-retest SRTs for either subgroup. Thus, the mean-SRT difference between the two subgroups was likely due to the higher number of participants with HL in the first subgroup, who also had poorer hearing thresholds (*p*= 0.001) compared to the second subgroup.

### Pure-tone hearing thresholds vs Remote-DIN test

Remote-DIN SRTs were also compared with lab-measured, better-ear PTA (BE-PTA _0.25, 0.5, 1, 2, 4, 8 kHz_) to examine sensitivity of the remote-DIN test as a measure of hearing loss (Fig. 4). Regression analysis showed that all 5 versions of the remote-DIN test significantly predicted BE-PTA (F_(5, 28)_ = 7.15, *p* = 0.0002) accounting for 48% of variance. After removing the Diotic SRT with 4 kHz filtered noise (VIF = 6.7), the remaining 4 DIN-SRTs significantly predicted BE-PTA (F_(4, 29)_ = 8.87, *p* < 0.0001) accounting for 49% of variance. Diotic SRT with 2 kHz filtered noise was the best predictor, explaining 50% of audiogram variance. There were significant correlations between all 5 versions of the remote-DIN and BE-PTA (*p* < 0.001 for total sample in each test). Diotic-BB was the only test version in which PTA and SRT were significantly correlated in both groups of participants (NH and HL). The slope of the Total regression line was also steeper in the diotic-BB than in any other version because the range and variability of SRT between participants was narrower.

**Figure 4.**
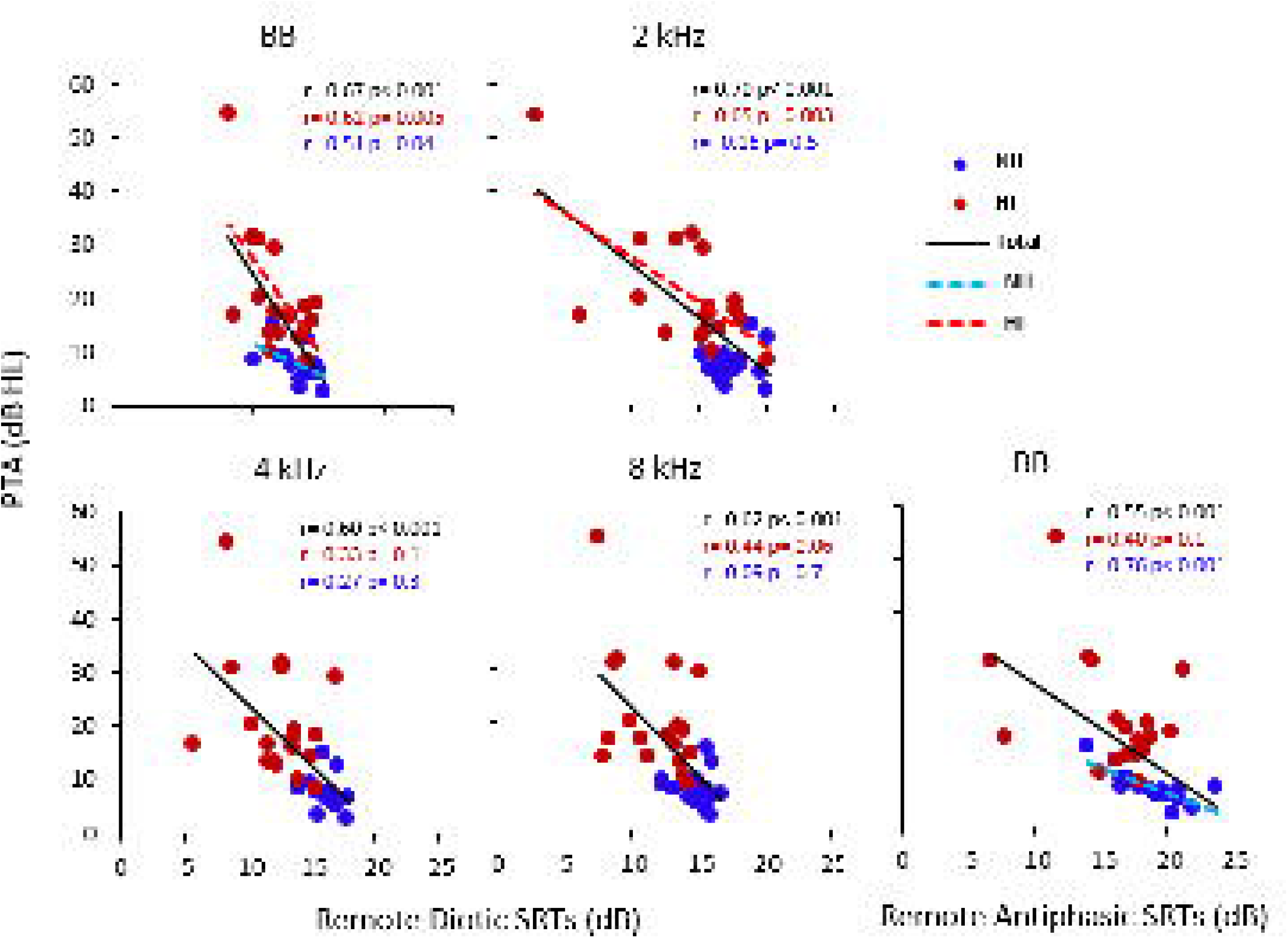
Remote DIN SRT predicted better-ear PTA. Details as in Fig. 3

There was one outlier in the HL group with moderate-severe hearing loss (Figure 4, PTA = 54 dB). Removing this outlier decreased the correlation between PTA and remote-DIN SRTs but did not change the overall significance of the relationship.

### SSQ-12 scores

SSQ scores had a broad range in both groups (Table 2a). A higher score indicated better perceived ability on the task. Overall, mean SSQ score was significantly higher (*p* = 0.03) in the NH than in the HL group. Among the 9 pragmatic subscales of SSQ (Noble et al, 2013), speech-in-speech, speech-in-noise, and quality and naturalness subscales had significantly higher mean scores in the NH than in the HL group (Table 2b).

**Table 2.** Descriptive statistics of SSQ scores in listeners with normal hearing (NH) and hearing loss (HL) grouped by **a)** overall score and **b)** subscale scores.

| **a) SSQ Overall Scores** |  |  |  |  |  |
| --- | --- | --- | --- | --- | --- |
| **Group** | **Mean** | **SD** | **Min** | **Max** | **P-value** |
| NH | 8.3 | 1.0 | 1.1 | 10 | 0.03 |
| HL | 7.6 | 1.4 | 1 | 10 |  |

| **b) SSQ Subscales** | **Group** | **Mean (SD)** | **P-value** |
| --- | --- | --- | --- |
| Speech in noise | NH | 8.6 (1.5) | 0.01* |
|  | HL | 7.4 (1.8) |  |
| Multiple speech streams | NH | 7.6 (2) | 0.2 |
|  | HL | 6.8 (2.1) |  |
| Speech in speech | NH | 8.2(1.8) | 0.02* |
|  | HL | 6.4 (2.4) |  |
| Localization | NH | 8.6 (1.4) | 0.7 |
|  | HL | 8.4 (2) |  |
| Distance and Movement | NH | 8.1 (1.8) | 0.8 |
|  | HL | 8.2 (2.1) |  |
| Segregation | NH | 8.6 (2.1) | 0.3 |
|  | HL | 8 (1.6) |  |
| Identification of sound | NH | 8.4 (1.4) | 0.6 |
|  | HL | 8.1(1.6) |  |
| Quality and naturalness | NH | 9.6 (0.5) | 0.03* |
|  | HL | 9 (0.9) |  |
| Listening effort | NH | 7.7 (2.2) | 0.1 |
|  | HL | 6.5 (2.3) |  |

### SSQ vs Remote-DIN test

There was a significant correlation between the remote DIN-SRTs and the SSQ scores across all listeners in all test versions, except the antiphasic BB-DIN test, where the relation was a non-significant trend (Fig. 5). Correlations were also significant in the HL group for the diotic BB and 4 and 8 kHz low-pass filtered test versions. The diotic BB version had the highest correlations and, again, the steepest regression slope.

**Figure 5.**
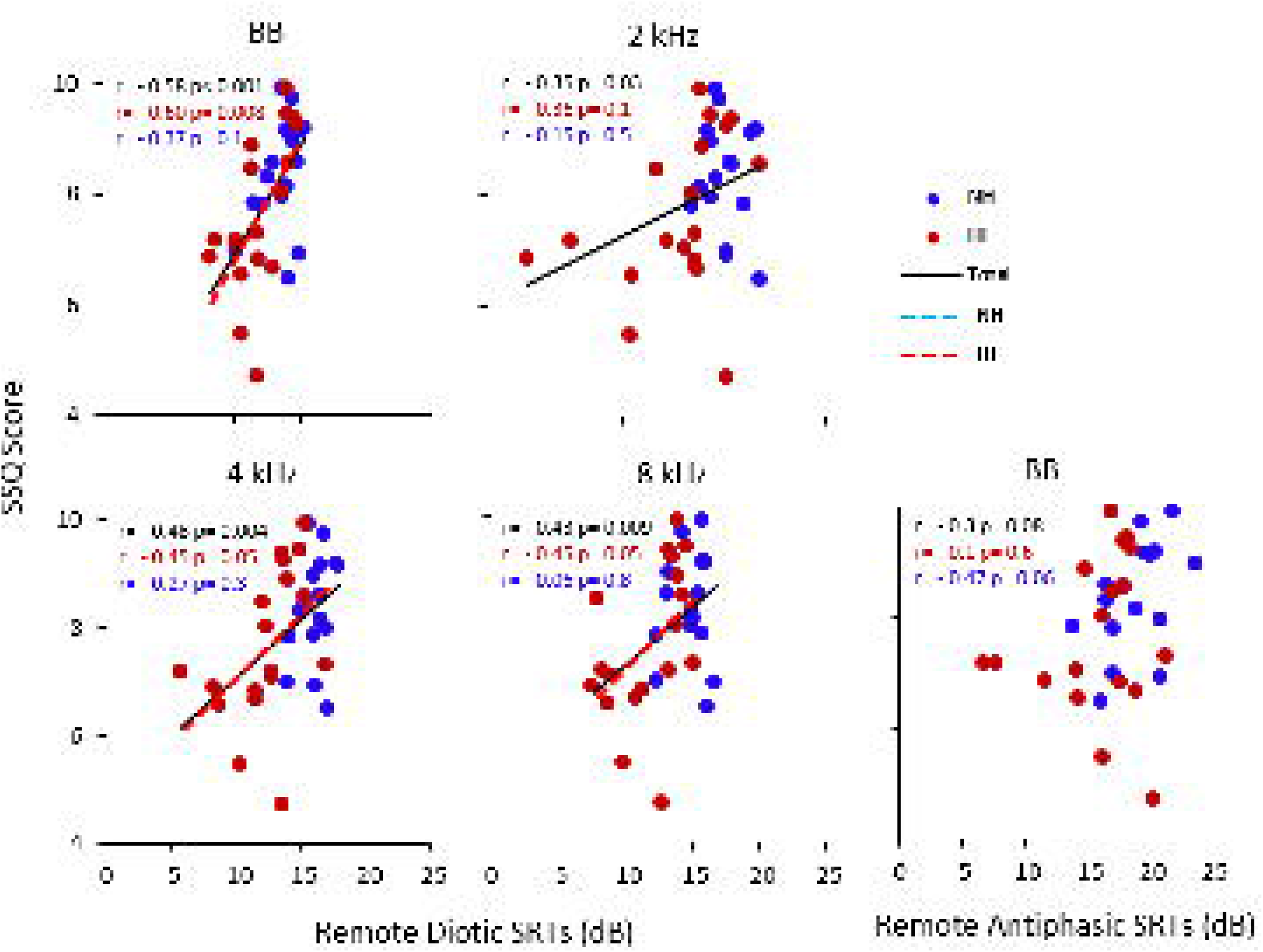
SSQ-12 predicted Remote DIN. Details as in Fig. 3

### SSQ vs Pure-tone hearing thresholds

SSQ mean score was a significant predictor of BE-PTA (Figure 6; F _(1, 32)_ =7.69, *p* = 0.009), independently accounting for 17% of variance in BE-PTA. Within group comparison showed the correlation was also significant for the NH group (Fig. 6). Significant correlations were found for both NH and HL groups, as well as the Total score (**NH:** r = - 0.63, *p* = 0.008; **HL:** r = - 0.48, *p* = 0.04; **Total:** r = -0.52, *p* = 0.001) when we compared SSQ scores only with <u>poorer ear</u> PTA.

**Figure 6.**
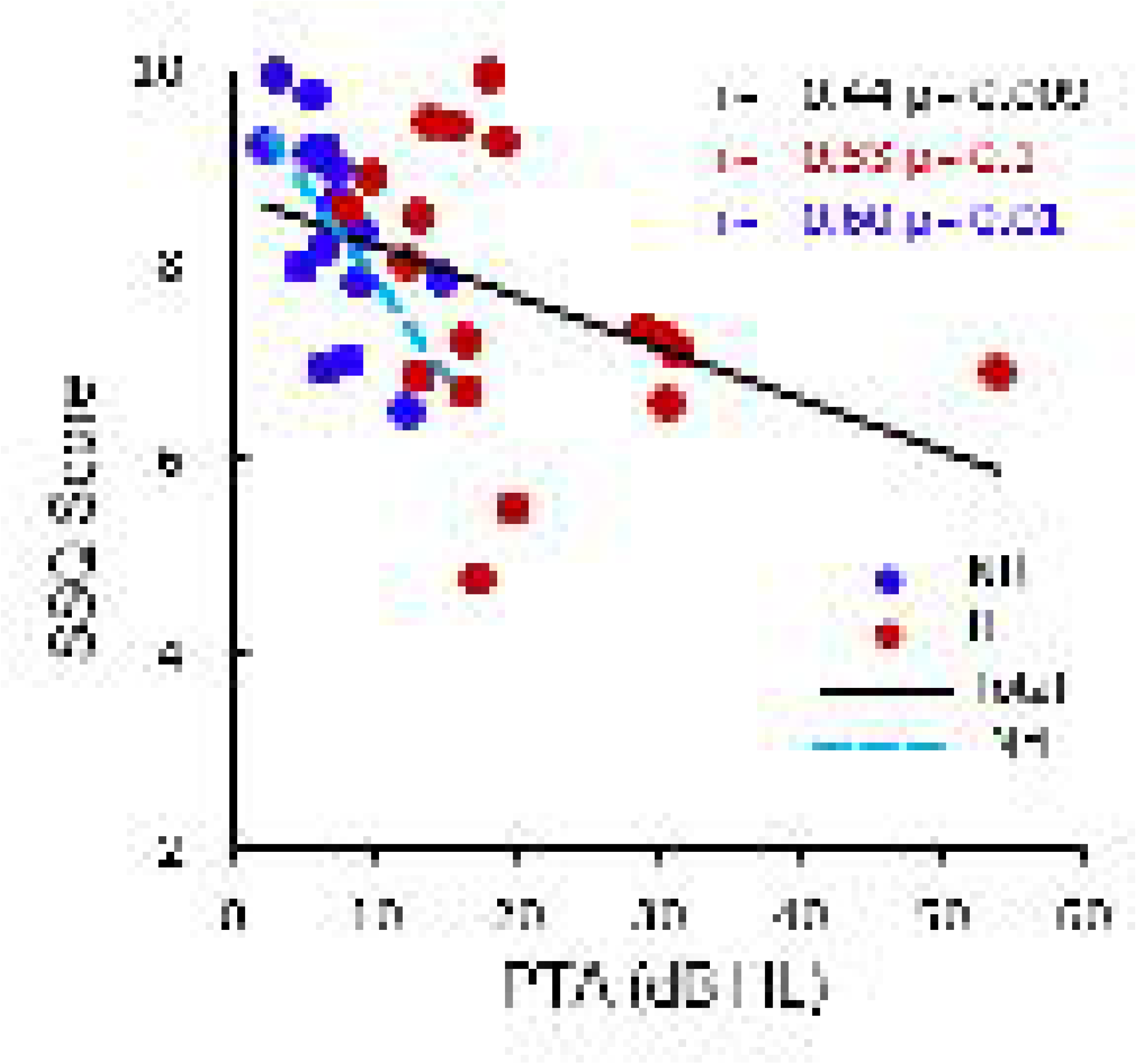
SSQ-12 predicted better-ear PTA. Details as in Fig. 3

## Discussion

The primary aims of this study were to examine the reliability of remote, self-administered DIN variants. Secondary aims were to examine predictive validity of the remote DIN and a self-report scale (SSQ-12) in comparison with each other and with pure-tone audiometry. Our results demonstrated the feasibility, reliability and validity of the DIN and SSQ-12 measures in a remote setting. Specifically, the DIN tests self-administered by our participants at home showed a strong correlation with the lab-version of the test, regardless of the types of headphones (or earbuds) and device (PC/smartphone/tablet) they used at home. All versions of the remote-DIN significantly predicted the audiogram, with the 2 kHz filtered version explaining 50% of audiogram variance. Current results were consistent with our previous studies (Motlagh Zadeh et al., 2020) showing lab-measured 2 kHz low-pass filtered DIN was the best version for detecting hearing loss between 0.25 to 4 kHz.

Self-report measures are inherently more variable than other, more objective measures because of their open-ended, interpretive nature. Despite compressed scoring ranges, SSQ predicted both SRTs and PTA, with speech-in-noise and sound quality subscales being the main predictors, rather than spatial hearing. The strong relationship between SSQ Overall Score and PTA in the NH group suggested that SSQ may be capturing sub-clinical aspects of HL.

All participants were able to perform and complete the remote tasks. Although the sample of participants was mostly young, college-graduate adults (64%; M= 34.2 y/o), 35% were older (> 50 y/o) and none of them reported any difficulty self-administering the DIN test at home. This is consistent with the study by Van der Mescht et al (2022) that assessed perception of cochlear implant users (18-78 y/o), in both home and clinical environments, using a 5-point Likert scale (from strongly agree to strongly disagree). All participants (N = 33) agreed or strongly agreed that they could perform the DIN test in both environments.

### Applications

The results have applied significance, especially during the Covid-19 pandemic that has moved audiology towards greater remote testing and accessibility, personalization and efficiency (Wasmann et al., 2021). Findings show that quantitative and qualitative remote testing can be used without sacrificing quality. Smartphone-mediated telehealth holds great promise to mitigate many of the barriers that prevent access to specialized hearing healthcare services (Swanepoel & Clark, 2019). Global availability of affordable internet, computers, and smart phones allow access to reliable, undemanding hearing assessment. These developments enable early identification and management of hearing loss, particularly for underserved communities, like rural populations and people who are unable to travel to receive in-person services (Swanepoel et al, 2019; De Sousa et al, 2020).

In addition to hearing loss detection, DIN tests have been shown to be useful for measuring the benefits of hearing devices, and for fitting hearing aids (Potgieter et al, 2018; Van der Mescht et al. 2022). Because they can be used at home, they can be helpful in monitoring hearing, for example for people with age-related hearing loss who have not yet chosen to use a hearing aid. With the development of drugs to protect hearing (Schilder et al, 2019), remote hearing assessment like DIN will be important both to detect candidates for drug administration and to monitor the effects of those drugs at home and on a regular basis. In addition, DIN testing has the potential to be used as a primary hearing screen in schools (for children 6+ y/o; Denys et al., 2018, 2021), and for patients who would benefit from frequent monitoring of ototoxicity, for example during aminoglycoside or cisplatin treatment.

Results of self-report (SSQ-12) were also significantly correlated with audiograms and DIN tests. These findings are consistent with our previous study (Motlagh Zadeh et al., 2019), showing that the extent of hearing loss was related to the number of individuals self-reporting difficulty hearing in noise based on a single question. Louw et al (2018) also showed that combining self-report with an audiometry screen increases test sensitivity (81.0%) for hearing loss, being most sensitive (86.1%) to identify high-frequency hearing loss. Moreover, Heinrich et al (2015) found that hearing-related self-report scales correlate significantly with different measures of speech perception in noise (e.g., DIN and sentence in noise test). These findings combined with our finding of speech-in-noise being the most significant subscale of SSQ-12 for distinguishing the HL group from the NH supports the importance of including both self-report and a speech-in-noise test in routine audiological assessment.

Perceived hearing difficulty in the presence of normal audiometric thresholds is a clinical challenge that is not currently being effectively addressed. Recently, Doherty and Singh (2020) used self-report trouble hearing in noise to measure benefits of a mild-gain hearing aid for middle-aged adults with clinically normal hearing. They found that, after wearing the hearing aid for 2 weeks, participants who previously self-reported difficulty hearing in noise had improved hearing handicap scores compared to those reporting no difficulty hearing in noise. Overall, linking audiometry and speech-in-noise testing to self-report measures of listening difficulties enhances our understanding of the listener experience in different circumstances and can also comment more widely on the usability and effectiveness of test procedures.

## Data Availability

The data that support the findings of this study are available on request from the corresponding author.

## Acknowledgements and Declaration of Participation and Conflicting Interests

This study was supported by NIH grant R21DC016241 and by the Cincinnati Children’s Hospital Research Foundation. David Moore receives support from the NIHR Manchester Biomedical Research Centre. L.M.Z., D.W.S and D.R.M. designed the experiments. L.M.Z. and V.B. collected the data. L.M.Z. and L.L. analyzed the data. L.M.Z., D.R.M and D.W.S. wrote the manuscript. D.W.S. and D.R.M. have a relationship with the hearX Group that includes equity, consulting, and potential royalties.

## References

American Academy of Otolaryngology–Head and Neck Surgery. AAO-HNS Bulletin/October 1993, 16–17.

American Academy of Otolaryngology–Head and Neck Surgery. AAO-HNS Bulletin/January 1994, 26–28.

Coco, L., Champlin, C. A., & Eikelboom, R. H. (2016). Community-Based Intervention Determines Tele-Audiology Site Candidacy. American journal of audiology, 25(3S), 264–267.

Davis, A., Smith, P., Ferguson, M., Stephens, D., & Gianopoulos, I. (2007). Acceptability, benefit and costs of early screening for hearing disability: A study of potential screening tests and models. Health Technology Assessment, 11, 1– 294.

De Sousa, K. C., Swanepoel, D. W., Moore, D. R., Smits, C. (2018). A smartphone national hearing test: performance and charectristics of users. American Journal of Audiology, 27, 448–454.

De Sousa, K. C., Smits, C., Moore, D. R., Myburgh, H. C., & Swanepoel, W. (2020). Pure-tone audiometry without bone-conduction thresholds: using the digits-in-noise test to detect conductive hearing loss. International journal of audiology, 59(10), 801–808.

De Sousa, K. C., Moore, D. R., Smits, C., & Swanepoel, D. W. (2021). Digital Technology for Remote Hearing Assessment—Current Status and Future Directions for Consumers. Sustainability, 13(18), 10124.

Denys, S., Hofmann, M., Luts, H., Guérin, C., Keymeulen, A., Van Hoeck, K., van Wieringen, A., Hoppenbrouwers, K., & Wouters, J. (2018). School-Age Hearing Screening Based on Speech-in-Noise Perception Using the Digit Triplet Test. Ear and hearing, 39(6), 1104–1115.

Denys, S., Wouters, J., & van Wieringen, A. (2021). The digit triplet test as a self-test for hearing screening at the age of school-entry. International journal of audiology, 1–8. Advance online publication.

Folmer, R. L., Vachhani, J., McMillan, G. P., Watson, C., Kidd, G. R., & Feeney, M. P. (2017). Validation of a computer-administered version of the digits-in-noise test for hearing screening in the United States. Journal of the American Academy of Audiology, 28(2), 161.

Gatehouse, S., & Noble, W. (2004). The speech, spatial and qualities of hearing scale (SSQ). International Journal of Audiology, 43(2), 85–99.

GSMA. The State of Mobile Internet Connectivity (2020). Retrieved from: https://www.gsma.com/r/wp-content/uploads/2020/09/GSMA-State-of-Mobile-Internet-Connectivity-Report-2020.pdf

Heinrich, A., Henshaw, H., & Ferguson, M. A. (2015). The relationship of speech intelligibility with hearing sensitivity, cognition, and perceived hearing difficulties varies for different speech perception tests. Frontiers in psychology, 6, 782.

Humes, L. E., Kinney, D. L., Main, A. K., & Rogers, S. E. (2019). A Follow-Up Clinical Trial Evaluating the Consumer-Decides Service Delivery Model. American journal of audiology, 28(1), 69–84. https://doi.org/10.1044/2018_AJA-18-0082

Karpa, M. J., Gopinath, B., Beath, K., Rochtchina, E., Cumming, R. G., Wang, J. J., & Mitchell, P. (2010). Associations between hearing impairment and mortality risk in older persons: The Blue Mountains Hearing Study. Annals of Epidemiology, 20, 452–459.

Leensen, M. C. J., de Laat, J. A, & Dreschler, W. A. (2011). Speech-in-noise screening tests by internet. Part 1: test evaluation for noise-induced hearing loss identification. International Journal of Audiology, 50, 823–34.

Louw, C., Swanepoel, W., & Eikelboom, R. H. (2018). Self-Reported Hearing Loss and Pure Tone Audiometry for Screening in Primary Health Care Clinics. Journal of primary care & community health, 9, 2150132718803156.

Maharani, A., Dawes, P., Nazroo, J., Tampubolon, G., Pendleton, N., & SENSE-Cog WP1 group (2018). Longitudinal Relationship Between Hearing Aid Use and Cognitive Function in Older Americans. Journal of the American Geriatrics Society, 66(6), 1130– 1136.

Moeller MP, Tomblin JB, Yoshinaga-Itano C, Connor CM, Jerger S. Current state of knowledge: Language and literacy of children with hearing impairment. Ear Hear 2007; 28:740–753.

Moore, D. R. (2018). Challenges in Diagnosing Auditory Processing Disorder. The Hearing Journal, 71 (10), 32–36.

Motlagh Zadeh, L., Silbert, H. N., Sternasty, K., Swanepoel, D. W., Hunter, L. L., Moore, D. R. (2019). Extended high frequency hearing enhances speech perception in noise. Proceedings of the National Academy of Sciences,116(47).

Motlagh Zadeh, L., Silbert, N. H., Swanepoel, W., & Moore, D. R. (2020). Improved Sensitivity of Digits-in-Noise Test to High-Frequency Hearing Loss. Ear and hearing, 42(3), 565–573.

Noble, W., Jensen, N. S., Naylor, G., Bhullar, N., & Akeroyd, M. A. (2013). A short form of the Speech, Spatial and Qualities of Hearing scale suitable for clinical use: the SSQ12. International journal of audiology, 52(6), 409–412.

Olusanya, B. O., Neumann, K. J., & Saunders, J. E. (2014). The global burden of disabling hearing impairment: a call to action. Bulletin of the World Health Organization, 92(5), 367–373.

Ozimek, E., Kutzner, D., Sęk, A., & Wicher, A. (2009). Development and evaluation of Polish digit triplet test for auditory screening. Speech Communication, 51, 307–316.

Potgieter, J. M., Swanepoel, d., Myburgh, H. C., Hopper, T. C., & Smits, C. (2016). Development and validation of a smartphone-based digits-in-noise hearing test in South African English. International journal of audiology, 55(7), 405–411.

Potgieter, J., Swanepoel, D. W., Myburgh, H. C., & Smits, C. (2018). The south african english smartphone digits-in-noise hearing test: Effect of age, hearing loss, and speaking competence. Ear and Hearing, 39(4), 656–663.

Powell, W., Jacobs, J. A., Noble, W., Bush, M. L., & Snell-Rood, C. (2019). Rural Adult Perspectives on Impact of Hearing Loss and Barriers to Care. Journal of community health, 44(4), 668–674.

Punch, R., Hyde, M., & Creed, P. A. (2004). Issues in the school-to-work transition of hard of hearing adolescents. American Annals of the Deaf, 149(1), 28–38.

Schilder, A. G. M., Su, M. P., Blackshaw, H., Lustig, L., Staecker, H., Lenarz, T., . . . Warnecke, A. (2019). Hearing protection, restoration, and regeneration: An overview of emerging therapeutics for inner ear and central hearing disorders. Otology & Neurotology, 40(5), 559–570.

Singh, J., & Doherty, K. A. (2020). Use of a Mild-Gain Hearing Aid by Middle-Age Normal-Hearing Adults Who Do and Do Not Self-Report Trouble Hearing in Background Noise. American journal of audiology, 29(3), 419–428.

Smits, C., Kapteyn, T. S., Houtgast, T. (2004). Development and validation of an automatic speech-in-noise screening test by telephone. International Journal of Audiology, 43, 15–28.

Smits, C., & Houtgast, T. (2005). Results from the Dutch speech-in-noise screening test by telephone. Ear and Hearing, 26, 89–95.

Smits, C., Kramer, S.E., & Houtgast, T. (2006). Speech reception thresholds in noise and self-reported hearing disability in a general adult population. Ear and Hearing, 27, 538–549.

Swanepoel, d., & Hall, J. W., 3rd (2010). A systematic review of telehealth applications in audiology. Telemedicine journal and e-health: the official journal of the American Telemedicine Association, 16(2), 181–200.

Swanepoel, D., & Clark, J. L. (2019). Hearing healthcare in remote or resource-constrained environments. The Journal of laryngology and otology, 133(1), 11–17.

Swanepoel, W., De Sousa, K. C., Smits, C., & Moore, D. R. (2019). Mobile applications to detect hearing impairment: opportunities and challenges. Bulletin of the World Health Organization, 97(10), 717–718.

Vlaming, Marcel S. M G, MacKinnon, R. C., Jansen, M., & Moore, D. R. (2014). Automated screening for high-frequency hearing loss. Ear and Hearing, 35, 667–679.

Van der Mescht L, le Roux T, Mahomed-Asmail F, De Sousa KC, Swanepoel D (2022). Remote monitoring of adult cochlear implant recipients using digits-in-noise self-testing. American Journal of Audiology, 31(3S), 923-935.

Warner-Czyz, A. D., Loy, B. A., Evans, C., Wetsel, A., & Tobey, E. A. (2015). Self-esteem in children and adolescents with hearing loss. Trends in Hearing, 19.

Wasmann, J. A., Lanting, C. P., Huinck, W. J., Mylanus, E. A. M., van der Laak, J. W. M., Govaerts, P. J., Swanepoel, W., Moore, D. R., & Barbour, D. L. (2021). Computational Audiology: New Approaches to Advance Hearing Health Care in the Digital Age. Ear and hearing, 42(6), 1499–1507. https://doi.org/10.1097/AUD.0000000000001041

World Health Organization. (2021). Deafness and Hearing Loss Factsheet. Retrieved from https://www.who.int/health-topics/hearing-loss#tab=tab_1

World Health Organization. (2021). Deafness and Hearing Loss. Retrieved from https://www.who.int/news-room/fact-sheets/detail/deafness-and-hearing-loss

